# Association of social media–sourced blood donors with transfusion delay and donor-related irregularities: A multicentre study in Bangladesh

**DOI:** 10.64898/2026.04.08.26350439

**Authors:** Ashraful Hoque, Marufur Rahman, Sushanta Kumar Basak, ABM Al Mamun

**Affiliations:** Department of Transfusion Medicine, Cumilla Medical College, Cumilla, Bangladesh; Senior Performance Measurement Specialist, Health System Performance Management, Ministry of Health, Government of Saskatchewan; Department of Transfusion Medicine, Faridpur Medical College, Bangladesh; Department of Transfusion Medicine, Dhaka Medical College, Bangladesh

**Keywords:** Blood donor systems, Transfusion delay, Social media, Hemovigilance, Low-resource settings, Blood safety

## Abstract

**Background:** In the absence of structured donor registries, social media platforms have become a dominant mechanism for blood donor recruitment in many low-resource settings. However, the implications of this shift for transfusion timeliness and system reliability remain unclear.

**Objective:** To evaluate the impact of social media–sourced donors on transfusion delay, donor reliability, and hemovigilance-related outcomes compared with conventional donor pathways.

**Methods:** This prospective analytical study included 400 transfusion episodes across tertiary hospitals in Bangladesh. Donor sources were categorized as social media (SM) or conventional (CON). The primary outcome was delay-to-transfusion. Secondary outcomes included donor-related irregularities, documentation completeness, near-miss events, and acute transfusion reactions. Multivariable logistic regression identified predictors of delay ≥4 hours.

**Results:** Social media–sourced donors were associated with significantly longer transfusion delays (5.98 vs 2.97 hours; p<0.001). Delay ≥4 hours occurred in 83.6% of SM cases versus 17.6% of CON cases (OR 23.78). Donor-related irregularities were observed in 85% of SM episodes and absent in CON donors. Safety outcomes did not differ significantly between groups. Social media donor sourcing remained the strongest independent predictor of delay (adjusted OR 18.09).

**Conclusion:** Unregulated social media–based donor recruitment introduces substantial delays and undermines system reliability without improving access. Integration of digital tools into regulated donor systems is essential to strengthen transfusion timeliness and hemovigilance in resource-limited settings.

## Introduction

Timely access to safe blood is not merely a logistical requirement but a critical determinant of survival in modern healthcare. From trauma resuscitation to obstetric hemorrhage and critical care, the effectiveness of transfusion therapy depends not only on compatibility and component quality but also on the speed with which blood becomes available [1–3]. Contemporary transfusion systems in high-income settings have evolved to address this need through centralized donor registries, coordinated blood services, and integrated digital infrastructure, ensuring both availability and traceability [4,5].

In contrast, many low- and middle-income countries (LMICs) continue to operate within fragmented transfusion ecosystems where structured donor pools are limited or absent. In such contexts, the responsibility of securing blood frequently shifts to patients’ families, leading to reliance on replacement donors and informal networks [6]. These systems are inherently unpredictable, often associated with delays, inconsistent donor reliability, and reduced oversight, all of which may compromise patient outcomes and weaken hemovigilance [7].

In Bangladesh, this structural gap has been increasingly filled by social media platforms, particularly Facebook, which function as de facto donor recruitment systems. These platforms enable rapid dissemination of requests and have created expansive, loosely organized donor communities. At face value, this represents a form of digital innovation—leveraging connectivity to address supply constraints. However, unlike formal digital donor registries implemented in regulated environments, social media–based recruitment operates without verification mechanisms, standardized workflows, or accountability structures.

This distinction is critical. Digital tools embedded within structured systems have demonstrated improvements in donor mobilization and response time [8,9]. Conversely, when digital platforms function outside regulatory frameworks, they may replicate—and potentially amplify—the inefficiencies of informal donor systems. Concerns include delayed donor arrival, high no-show rates, financial transactions, involvement of intermediaries, and incomplete documentation, all of which may affect both timeliness and safety.

Timeliness, in particular, is a clinically sensitive endpoint. Evidence from trauma and hemorrhage literature consistently demonstrates that early transfusion is associated with improved survival, whereas delays contribute to worse outcomes [2,10]. In settings where transfusion is urgently required, even modest delays can translate into significant clinical consequences.

Despite the widespread adoption of social media for donor recruitment in Bangladesh and similar settings, there is a notable absence of empirical data evaluating its impact on transfusion processes. Existing literature has largely focused on donor motivation and awareness rather than operational performance or system-level outcomes.

This study addresses that gap by systematically comparing social media–sourced donors with conventional donor pathways across multiple centers. By examining transfusion delay, donor-related irregularities, and safety outcomes, the study aims to provide evidence that informs both clinical practice and policy development, particularly in the context of evolving digital health ecosystems in transfusion medicine.

## Methods

### Study Design and Setting

A prospective analytical cross-sectional study was conducted across tertiary-level hospitals in Bangladesh, including centers in Dhaka, Chattogram, Cumilla, and Faridpur.

### Study Population

A total of **400 transfusion episodes** were included.

### Exposure Groups

- **Social Media (SM):** Donors recruited via online platforms
- **Conventional (CON):** Donors from family, friends, hospital lists, or walk-in

### Data Variables

- Patient demographics and clinical urgency
- Donor pathway and fraud indicators
- Time intervals (request → donor arrival → transfusion)
- utcomes: delay ≥4h, reactions, near-miss events

### Statistical Analysis

- Continuous variables: Welch’s t-test
- Categorical variables: Chi-square/Fisher’s exact test
- Logistic regression for predictors of delay ≥4 hours
- Significance level: p < 0.05

## Results

### Study Population and Baseline Characteristics

A total of 400 transfusion episodes were prospectively enrolled across two tertiary-level hospitals in Bangladesh. Of these, 173 (43.3%) involved donors recruited through social media platforms (SM group) and 227 (56.8%) involved conventional donors sourced through family, friends, hospital lists, etc. (CON group). Baseline demographic and clinical characteristics are presented in **Table 1**. The two groups were comparable in age (SM: 41.2 ± 13.9 years; CON: 41.4 ± 13.9 years; *p-value* = 0.89), sex distribution (SM: 46.2% female; CON: 48.0% female; *p-value* = 0.72), clinical urgency (*p-value* = 0.33), transfusion indication (*p-value* = 0.37), blood component type (*p-value* = 0.68), and number of requested units (*p-value* = 0.48).

**Table 1.** Baseline demographic and clinical characteristics of 400 transfusion episodes by donor source.

| Characteristic | Social Media (n = 173) | Conventional (n = 227) | p-value |
| --- | --- | --- | --- |
| Age, years, mean ± SD | 41.2 ± 13.9 | 41.4 ± 13.9 | 0.89 |
| Female sex, n (%) | 80 (46.2) | 109 (48.0) | 0.72 |
| **Clinical urgency, n (%)** |  |  | 0.33 |
| Emergency | 70 (40.5) | 88 (38.8) |  |
| Urgent | 73 (42.2) | 88 (38.8) |  |
| Elective | 30 (17.3) | 51 (22.5) |  |
| **Transfusion indication, n (%)** |  |  | 0.37 |
| Surgery / Trauma | 31 (17.9) | 51 (22.5) |  |
| Pediatric | 32 (18.5) | 50 (22.0) |  |
| Hematology / Oncology | 34 (19.7) | 47 (20.7) |  |
| Obstetric | 36 (20.8) | 40 (17.6) |  |
| Medical Anaemia | 40 (23.1) | 39 (17.2) |  |
| **Blood component, n (%)** |  |  | 0.68 |
| Whole Blood | 50 (28.9) | 67 (29.5) |  |
| Packed Red Cells | 39 (22.5) | 51 (22.5) |  |
| Platelets | 48 (27.7) | 53 (23.3) |  |
| Plasma | 36 (20.8) | 56 (24.7) |  |
| **Ward type, n (%)** |  |  | 0.10 |
| Emergency | 45 (26.0) | 53 (23.3) |  |
| General Ward | 44 (25.4) | 63 (27.8) |  |
| Operation Theatre | 34 (19.7) | 65 (28.6) |  |
| ICU / HDU | 50 (28.9) | 46 (20.3) |  |
| Requested units, mean ± SD | 1.99 ± 0.84 | 1.93 ± 0.84 | 0.48 |

*Continuous variables expressed as mean ± SD; categorical variables as n (%). p-values from Welch's t-test (continuous) or Chi-square test (categorical). SM = social media; CON = conventional.*

Among SM-sourced donors, 33 (19.1%) did not attend despite initial commitment (donor no-show), exclusively within the social media group. For analyses involving delay-to-transfusion and clinical outcomes (transfusion reactions, near-miss events), these 33 episodes were excluded, yielding a working sample of 367 episodes (SM: 140; CON: 227). The rationale was that downstream variables could not be attributed to a donor who never arrived. Donor no-show was retained as a fraud indicator in the full-sample analysis (N = 400).

### Primary Outcome: Delay to Transfusion

Social media-sourced donors were associated with significantly longer delay-to-transfusion compared with conventional donors (**Figure 1A**). The mean delay was 5.98 hours (SD 2.02) in the SM group versus 2.97 hours (SD 0.97) in the CON group, yielding a mean difference of 3.02 hours (95% CI: 2.66–3.38; *t* = 16.52, df = 179.3, *p* < 0.0001, Welch’s *t*-test). The Shapiro-Wilk test confirmed normality in both groups (SM: W = 0.997, *p* = 0.96; CON: W = 0.991, *p* = 0.16). The effect size was large (η^2^ = 0.604), indicating that donor source alone explained approximately 60% of the variance in transfusion delay.

**Figure 1.**
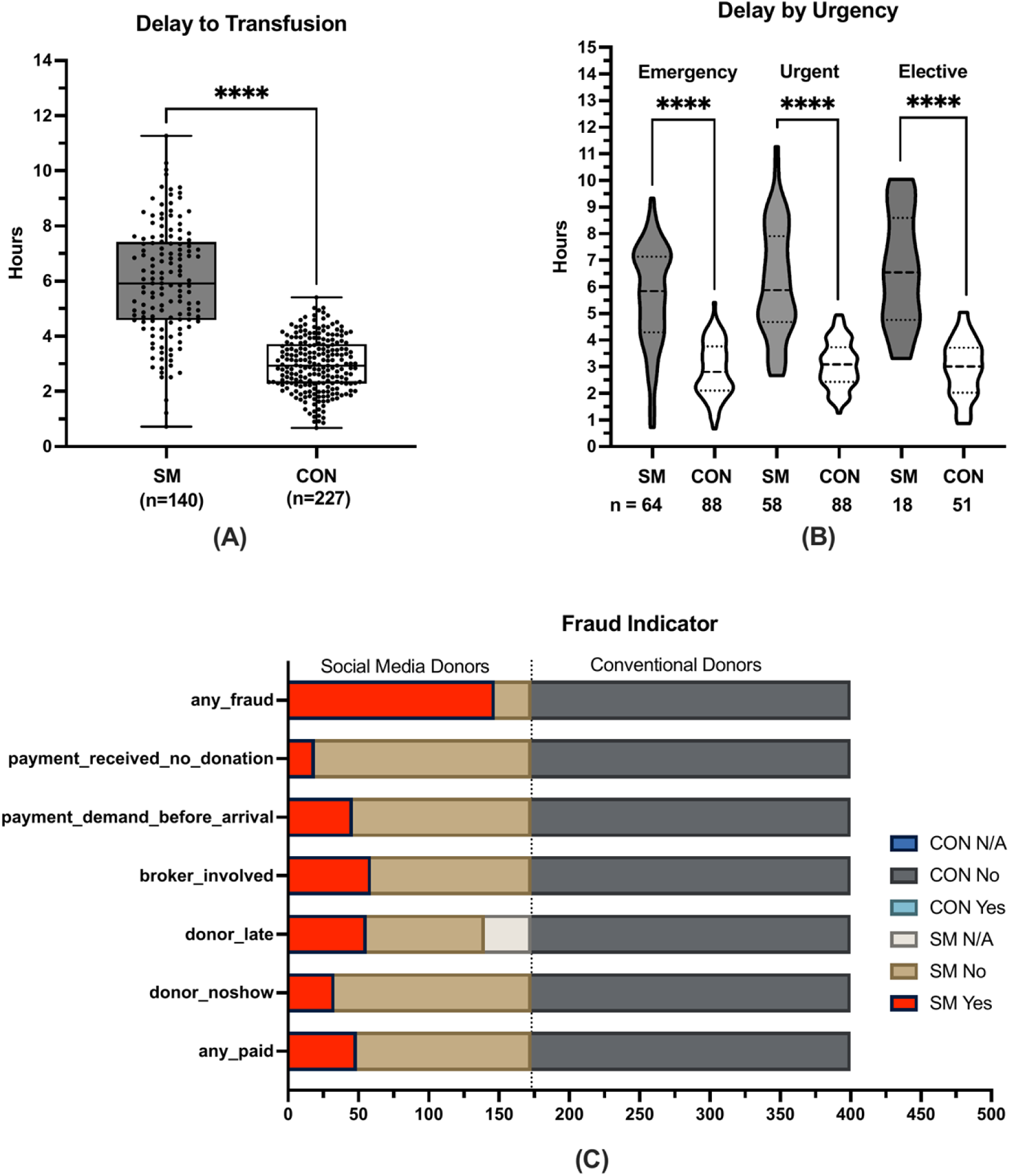
Delay to transfusion and fraud indicators by donor source. **(A)** Box-and-whisker plot of delay to transfusion (hours) for social media (SM, n = 140) versus conventional (CON, n = 227) donors (excluding no-shows). Boxes represent interquartile range; whiskers represent range. \*\*\*\**p* < 0.0001, Welch’s *t*-test. **(B)** Delay to transfusion stratified by clinical urgency. The SM–CON difference was significant within all three urgency subgroups (all *p-value* < 0.0001). **(C)** Stacked bar chart of fraud indicators by donor source. All six fraud indicators occurred exclusively in the SM group (all *p-value* < 0.0001, Fisher’s exact test). N/A denotes cases where the variable was set to missing due to donor no-show.

When dichotomised at the clinically meaningful threshold of 4 hours, 83.6% (117/140) of SM-sourced transfusion episodes experienced a delay ≥4 hours, compared with only 17.6% (40/227) of conventional episodes (OR = 23.78; 95% CI: 13.48–40.57; *p-value* < 0.0001, Fisher’s exact test).

#### Subgroup Analysis: Delay by Clinical Urgency

The delay disparity between SM and CON donors was consistent across all urgency categories (**Figure 1B**). In the emergency subgroup, SM donors had a mean delay of 5.64 hours versus 2.88 hours for CON donors (mean difference: 2.76 hours; 95% CI: 2.24–3.28; *t* = 10.61, df = 89.2, *p* < 0.0001; n = 64 vs 88). In the urgent subgroup, SM donors had a mean delay of 6.17 hours versus 3.09 hours (mean difference: 3.08 hours; 95% CI: 2.51–3.65; *t* = 10.69, df = 70.1, *p-value* < 0.0001; n = 58 vs 88). In the elective subgroup, SM donors had a mean delay of 6.63 hours versus 2.93 hours (mean difference: 3.71 hours; 95% CI: 2.61–4.80; *t* = 7.05, df = 20.1, *p-value* < 0.0001; n = 18 vs 51). The largest absolute difference was observed in elective cases (η^2^ = 0.712), though the effect was substantial across all categories.

#### Fraud and Misconduct Indicators

Fraudulent or irregular donor behaviours were observed exclusively in the social media group (**Table 2**; **Figure 1C**). Among 173 SM-sourced episodes, 85.0% (147/173) exhibited at least one fraud indicator, compared with 0% in the conventional group (*p* < 0.0001). The most frequent individual indicators in the SM group were: donor late arrival (40.0%, 56/140 excluding no-shows), broker or middleman involvement (34.1%, 59/173), payment demand (28.3%, 49/173), pre-arrival payment demand (26.6%, 46/173), donor no-show (19.1%, 33/173), and payment received without donation (11.0%, 19/173). All indicators showed *p-value* < 0.0001 by Fisher’s exact test.

**Table 2.**
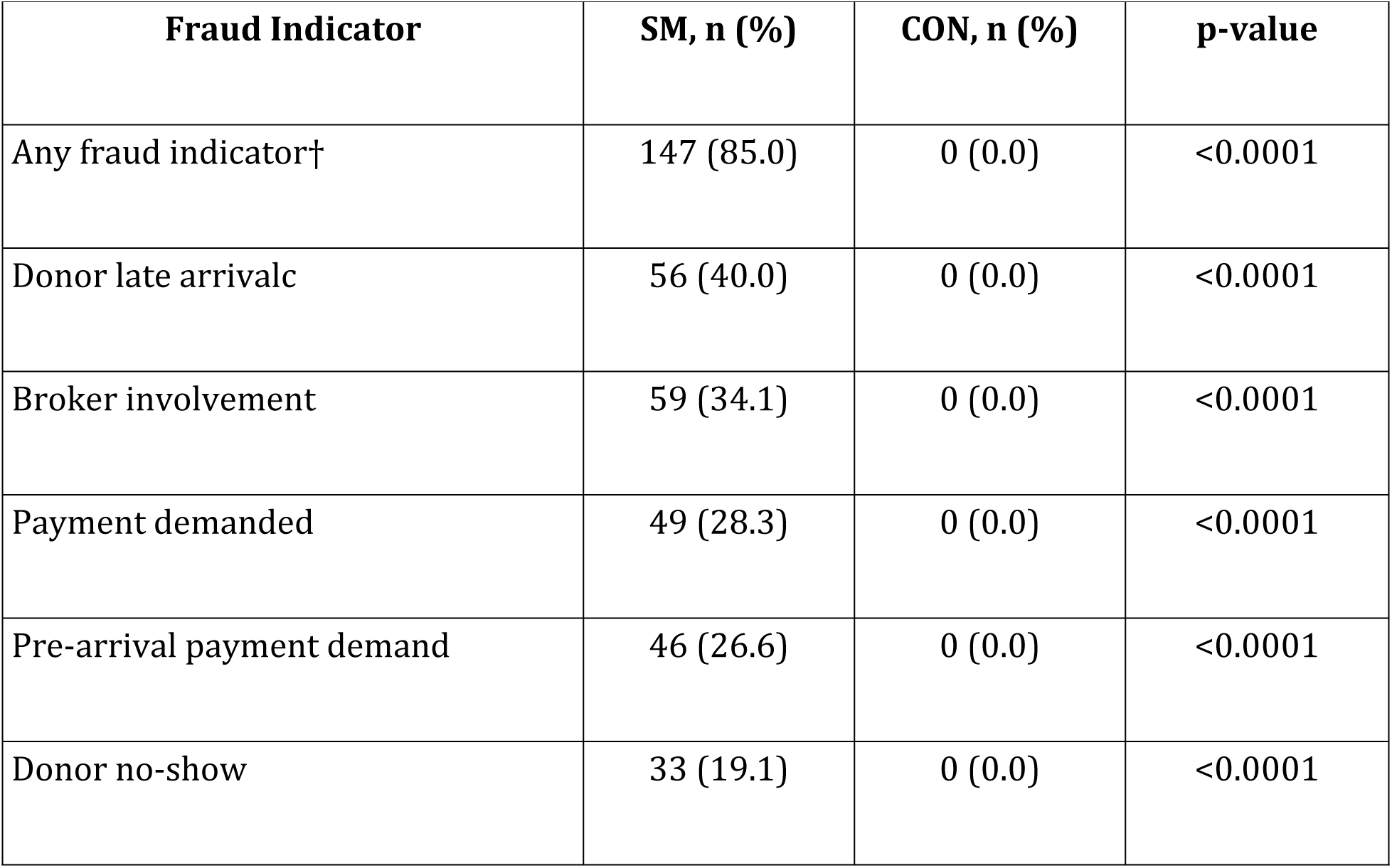
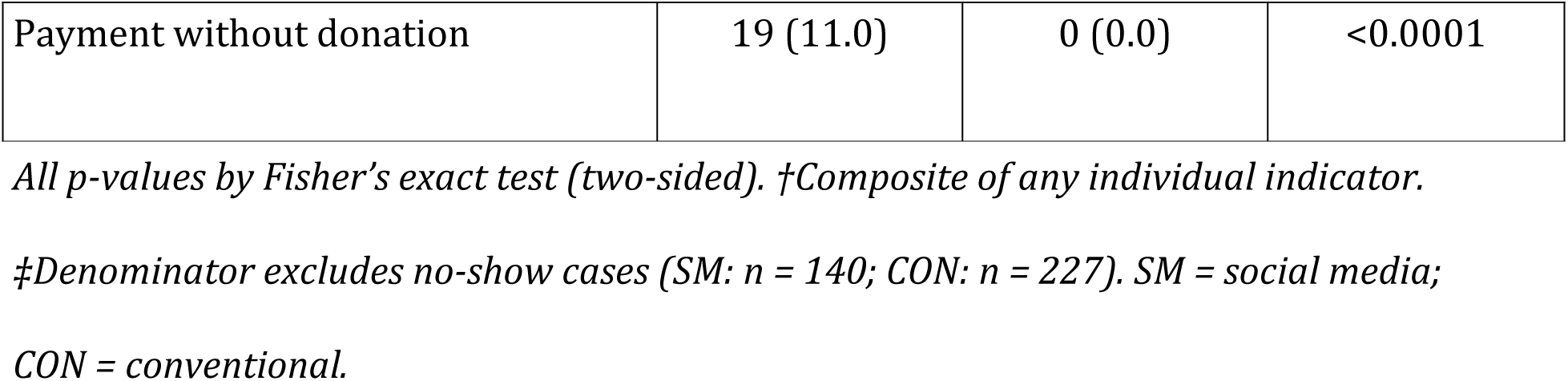
Prevalence of fraud and misconduct indicators by donor source (N = 400).

#### Documentation Completeness and Transfusion Safety

Documentation completeness and acute safety outcomes are summarised in **Table 3**. Documentation completeness did not differ significantly between groups: 72.3% (125/173) of SM episodes and 65.6% (149/227) of CON episodes had complete screening and crossmatch documentation (OR = 1.36; 95% CI: 0.89–2.08; *p* = 0.19).

**Table 3.** Documentation completeness and acute transfusion safety outcomes by donor source.

| Outcome | SM, n (%) | CON, n (%) | OR (95% CI) | p-value |
| --- | --- | --- | --- | --- |
| Documentation complete† | 125/173<br>(72.3) | 149/227<br>(65.6) | 1.36 (0.89–2.08) | 0.19 |
| Reaction within 24 h‡ | 20/140 (14.3) | 41/227 (18.1) | 0.76 (0.42–1.35) | 0.39 |
| Near-miss event‡ | 10/140 (7.1) | 17/227 (7.5) | 0.95 (0.44–2.15) | >0.99 |

*p-values by Fisher's exact test (two-sided). OR = odds ratio; CI = confidence interval.*
*†Full sample (N = 400). ‡Excludes donor no-show cases (N = 367; SM: 140, CON: 227).*
*SM = social media; CON = conventional.*

Among the 367 episodes where transfusion occurred (excluding no-shows), acute transfusion reactions within 24 hours were reported in 14.3% (20/140) of SM episodes versus 18.1% (41/227) of CON episodes (OR = 0.76; 95% CI: 0.42–1.35; *p-value* = 0.39). Near-miss events were similarly uncommon: 7.1% (10/140) in the SM group versus 7.5% (17/227) in the CON group (OR = 0.95; 95% CI: 0.44–2.15; *p-value* > 0.99). Neither outcome differed significantly by donor source, suggesting that once a compatible unit was obtained, the acute clinical safety profile was comparable.

#### Multivariable Logistic Regression: Predictors of Delay ≥4 Hours

Multivariable logistic regression was performed to identify independent predictors of clinically significant delay (≥4 hours) among the 367 episodes where donors attended (**Table 4**; **Figure 2**). After adjusting for age, sex, urgency category, number of requested units, payment demand, broker involvement, and documentation completeness, social media donor source remained the strongest independent predictor of delay (adjusted OR = 18.09; 95% CI: 9.26–37.36; *p-value* < 0.001). No other covariates achieved statistical significance in the adjusted model.

**Table 4.** Univariable and multivariable logistic regression: predictors of transfusion delay ≥4 hours (N = 367, excluding donor no-shows).

| **Variable** | **Unadjusted** |  |  | **Adjusted** |  |  |
| --- | --- | --- | --- | --- | --- | --- |
|  | **OR** | **95% CI** | **p** | **aOR** | **95% CI** | **p** |
| SM donor (vs CON) | 23.78 | 13.79–42.59 | <0.001 | 18.09 | 9.26–37.36 | <0.001 |
| Age (per year) | 0.99 | 0.97–1.00 | 0.158 | 0.98 | 0.96–1.00 | 0.098 |
| Female (vs male) | 1.23 | 0.82–1.87 | 0.320 | 1.49 | 0.85–2.62 | 0.162 |
| Emergency (vs elective) | 1.05 | 0.69–1.59 | 0.835 | 0.81 | 0.38–1.77 | 0.602 |
| Urgent (vs elective) | 1.18 | 0.77–1.80 | 0.445 | 1.02 | 0.47–2.24 | 0.954 |
| Requested units | 1.07 | 0.83–1.37 | 0.615 | 1.05 | 0.75–1.46 | 0.773 |
| Payment demanded | 15.88 | 6.18–53.99 | <0.001 | 2.44 | 0.82–9.01 | 0.135 |
| Broker involved | 11.24 | 4.96–30.26 | <0.001 | 1.51 | 0.56–4.57 | 0.432 |
| Documentation complete | 1.44 | 0.92–2.28 | 0.111 | 1.31 | 0.72–2.40 | 0.380 |

*OR = unadjusted odds ratio; aOR = adjusted odds ratio; CI = confidence interval.*
*Reference categories: conventional donor, male sex, elective urgency. Model AUC (C-statistic) = 0.859; classification accuracy = 82.8%. SM = social media; CON = conventional.*

**Figure 2.**
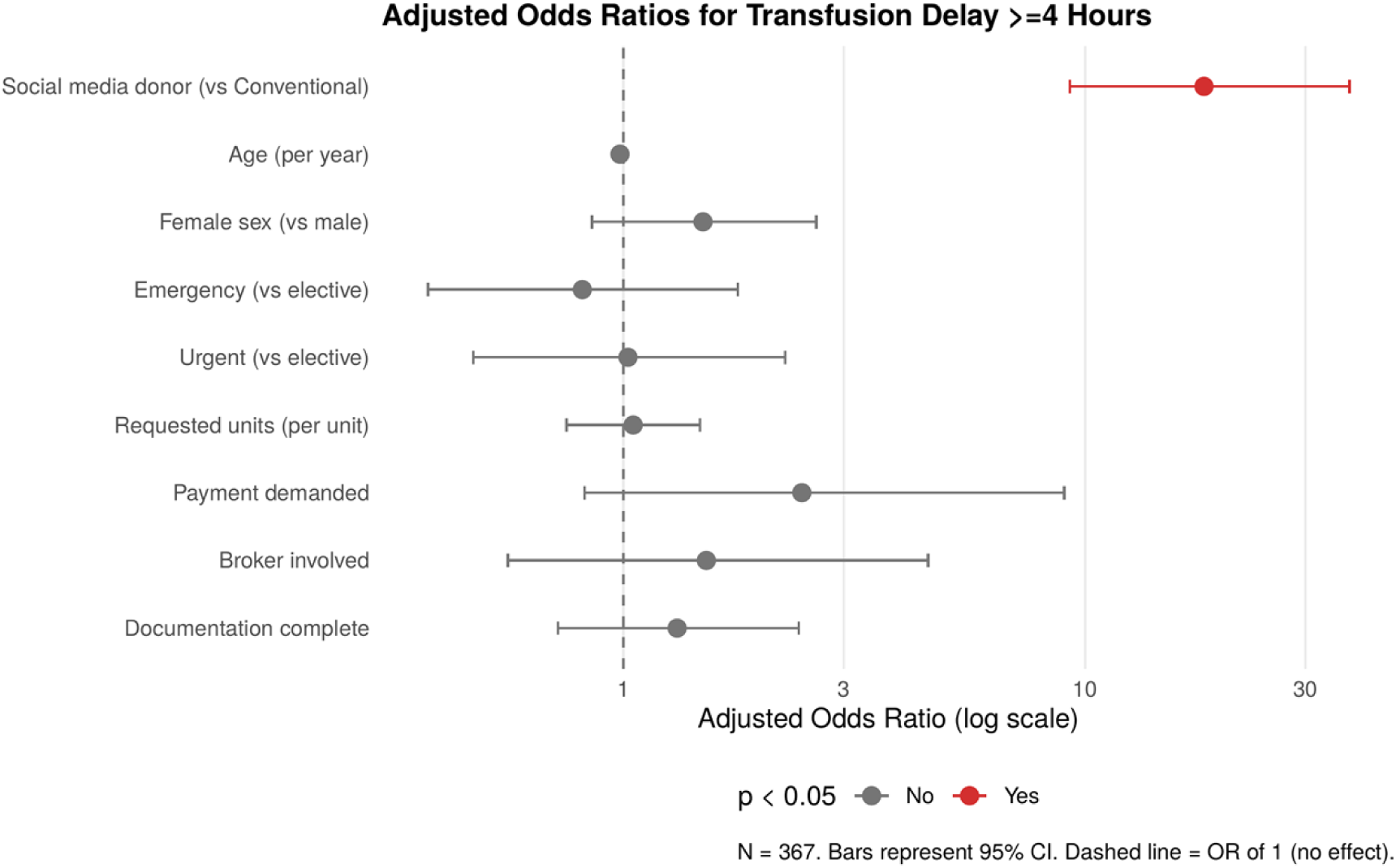
Forest plot of adjusted odds ratios (aOR) for transfusion delay ≥4 hours. Points represent aOR estimates; horizontal bars represent 95% confidence intervals. The dashed vertical line indicates OR = 1 (null effect). Red points denote statistically significant predictors (*p* < 0.05); grey points denote non-significant predictors. Social media donor source was the only significant independent predictor (aOR = 18.09; 95% CI: 9.26–37.36). N = 367.

Notably, payment demand (unadjusted OR = 15.88; 95% CI: 6.18–53.99; *p* < 0.001) and broker involvement (unadjusted OR = 11.24; 95% CI: 4.96–30.26; *p-value* < 0.001) were strongly associated with delay in univariable analysis but were attenuated and non-significant after adjustment (aOR = 2.44, *p-value* = 0.14; and adjusted-OR = 1.51, *p* = 0.43, respectively). This attenuation suggests that these fraud-related behaviours are mediators of the social media pathway rather than independent risk factors, and that the social media donor source variable captures the aggregate risk introduced by these mechanisms.

The model demonstrated good discriminatory performance, with an area under the receiver operating characteristic curve (AUC) of 0.859 and an overall classification accuracy of 82.8%.

### Summary of Key Findings

In summary, social media-sourced donors were associated with: (1) a threefold longer mean delay to transfusion (5.98 vs 2.97 hours); (2) a 24-fold higher odds of clinically significant delay ≥4 hours (OR = 23.78); (3) near-universal prevalence of at least one fraud indicator (85.0% vs 0%); and (4) no significant differences in acute transfusion reactions or near-miss events once transfusion was completed. The delay effect persisted across all urgency subgroups and was robust to multivariable adjustment. Social media donor source was the single dominant independent predictor of delay in logistic regression (aOR = 18.09; AUC = 0.859).

## Discussion

This study demonstrates that social media–based donor recruitment, although widely adopted as a pragmatic solution in Bangladesh, is associated with substantial delays in transfusion and a high burden of donor-related irregularities. These findings highlight a fundamental tension between **accessibility and system reliability** in contemporary transfusion practice within resource-limited settings.

The magnitude and consistency of transfusion delay observed in this study are particularly noteworthy. Social media–sourced donors were associated with an approximately three-hour increase in time to transfusion, a difference that persisted across emergency, urgent, and elective scenarios. In transfusion medicine, such delays are not trivial. Early administration of blood components has been shown to be critical in trauma resuscitation and hemorrhagic shock, where delays are linked to increased morbidity and mortality [2,10]. The persistence of delay across clinical contexts suggests that the underlying issue is not related to prioritization of care but rather reflects intrinsic inefficiencies within the donor acquisition pathway.

The multivariable analysis reinforces this interpretation. Social media donor sourcing remained the dominant independent predictor of delay, even after adjusting for clinical and operational variables. The attenuation of individual fraud-related factors in the adjusted model suggests that these behaviors are not independent drivers but rather manifestations of a broader, unregulated system. In effect, social media donor sourcing encapsulates a cluster of structural vulnerabilities, including uncertainty of donor commitment, lack of verification, and the presence of intermediaries.

The high prevalence of donor-related irregularities further underscores the systemic nature of the problem. Behaviors such as late arrival, payment demands, broker involvement, and donor no-show were confined exclusively to the social media group. These findings are consistent with previous observations in settings where informal or replacement donor systems predominate, often leading to commercialization and reduced accountability [6,11]. Importantly, these practices not only delay transfusion but also shift the burden of risk—financial, emotional, and clinical—onto patients and their families.

A key insight from this study is that transfusion safety outcomes, including acute reactions and near-miss events, did not differ significantly between groups. This suggests that once a compatible unit is secured and enters the formal healthcare system, existing laboratory and clinical safeguards function effectively. Therefore, the primary vulnerability lies upstream, at the interface between donor identification and blood collection. This distinction is critical for policy and system design, indicating that improving transfusion outcomes in such settings requires strengthening donor recruitment systems rather than focusing solely on laboratory processes.

From a broader perspective, these findings reflect the evolving role of digital platforms in healthcare delivery. Social media represents a powerful tool for communication and community mobilization, but its effectiveness in clinical systems depends on integration with structured processes. In high-income settings, digital donor registries are embedded within regulated frameworks that ensure donor eligibility, traceability, and timely mobilization [4,8]. In contrast, the unregulated use of social media observed in this study lacks these safeguards, resulting in variability and unpredictability.

The implications for hemovigilance are also significant. Effective hemovigilance relies on traceability and accountability across the transfusion chain. Informal donor pathways may weaken this chain, particularly when intermediaries are involved or when documentation is incomplete. Although documentation completeness did not differ significantly in this study, the presence of informal practices raises concerns about hidden gaps that may not be captured in routine data.

This study has several limitations. It was conducted in tertiary care settings and may not fully represent peripheral healthcare facilities. Fraud indicators were operationally defined and may be subject to reporting variability. Additionally, the study was not designed to assess long-term clinical outcomes such as mortality. However, the strength and consistency of the observed associations, along with the multicentre design, support the validity and generalizability of the findings within similar healthcare contexts.

Overall, the study highlights that while social media expands the reach of donor recruitment, it does not inherently improve system performance. Without integration into regulated frameworks, such platforms may introduce inefficiencies that undermine the timeliness and reliability of transfusion services.

## Conclusion

The increasing reliance on social media for blood donor recruitment in Bangladesh reflects both innovation and systemic necessity. However, this study demonstrates that such reliance is associated with significant delays in transfusion and a high burden of donor-related irregularities, without measurable improvement in clinical safety outcomes.

These findings emphasize that **availability alone is insufficient**; reliability and timeliness are equally critical components of an effective transfusion system. Social media, in its current unregulated form, does not meet these requirements.

Moving forward, the focus should not be on replacing digital platforms but on redefining their role within structured transfusion systems. Integrating social media–based outreach into verified donor registries, strengthening hospital-based donor pools, and implementing digital verification and tracking mechanisms are essential steps toward improving system performance.

In resource-limited settings, where delays can directly translate into preventable morbidity and mortality, ensuring timely and reliable access to blood is a critical priority. Aligning technological innovation with system governance will be key to achieving this goal.

## “WHAT THIS STUDY ADDS”

### What is known

- Structured donor systems improve transfusion timeliness and safety
- Replacement and informal donor systems are associated with inefficiencies
- Digital tools can enhance donor recruitment when integrated into regulated systems

### What this study shows

- Social media–based donor recruitment significantly delays transfusion (≈3 hours)
- Donor-related irregularities (fraud, no-show, payment demand) are highly prevalent
- Social media donor sourcing is the strongest independent predictor of delay

### What this study adds

- First real-world, multicentre evidence quantifying the operational impact of social media donor systems
- Demonstrates that unregulated digital recruitment may undermine system reliability
- Highlights the need to integrate digital platforms into structured, verified donor systems

## AUTHOR CONTRIBUTIONS (CRediT TAXONOMY)

**Conceptualization:** Ashraful Hoque

**Methodology:** Ashraful Hoque, Marufur Rahman

**Data Curation:** Sushanta Kumar Basak, ABM Al Mamun

**Formal Analysis:** Marufur Rahman (statistician)

**Investigation:** Ashraful Hoque, Marufur Rahman, Sushanta Kumar Basak, ABM Al Mamun

**Writing – Original Draft:** Ashraful Hoque

**Writing – Review & Editing:** Ashraful Hoque, Marufur Rahman, Sushanta Kumar Basak, ABM Al Mamun

**Supervision:** Ashraful Hoque

## ETHICS STATEMENT

This study was conducted in accordance with the Declaration of Helsinki. Ethical approval was obtained from the appropriate institutional review boards. As this was an observational study using routine clinical data, informed consent requirements were waived where applicable.

## DATA AVAILABILITY STATEMENT

The dataset generated and analyzed during the current study is available from the corresponding author upon reasonable request. De-identified data may be shared in accordance with institutional and ethical guidelines.

